# Hemoglobin-to-Red Cell Distribution Width Ratio Predicts Three-Year Major Adverse Cardiovascular Events After Percutaneous Coronary Intervention: A Vietnamese Multicenter Cohort Study

**DOI:** 10.64898/2026.07.25.26358909

**Authors:** Hai Nguyen Ngoc Dang, Thang Viet Luong, Thanh Thien Tran, Thanh Van Ho, Mai Thi Thu Cao, Tan Ngoc Nguyen, Khiem Dong Thien, Cuong Hai Nguyen, Hung Minh Nguyen, Binh Anh Ho, Tien Anh Hoang, Minh Van Huynh

## Abstract

**Introduction:** Percutaneous coronary intervention (PCI) is a widely adopted strategy for managing coronary artery disease (CAD), leading to improved survival rates, particularly in developing countries. However, the increased survival of these patients imposes a substantial burden on long-term management, especially at the primary healthcare level. Recently, the hemoglobin-to-red cell distribution width ratio (HRR) has emerged as a potentially valuable prognostic biomarker for post-PCI patients. Despite its accessibility and cost-effectiveness, HRR has not been extensively investigated in resource-limited settings.

**Aim:** This study aimed to evaluate the prognostic value of the HRR in predicting 3-year major adverse cardiovascular events (MACE) among patients undergoing PCI who were managed at the primary healthcare level.

**Methods:** We conducted a multicenter prospective cohort study in Vietnam. A total of 626 post-PCI patients were ultimately included in the final analysis. The study commenced in October 2019 and concluded in October 2025. The association between HRR and 3-year MACE was evaluated using Cox proportional hazards regression models.

**Results:** The MACE incidence decreased progressively across the ascending HRR quartiles (p < 0.001). According to the unadjusted Cox model, each unit increase in HRR was associated with a lower risk of MACE (HR = 0.756; 95% CI, 0.703–0.814; p < 0.001). This association persisted after adjustment for age, sex, comorbidities (Model I: HR = 0.810; 95%CI: 0.750–0.878; p < 0.001). HRR outperformed its individual components, hemoglobin and red cell distribution width. Subgroup analyses confirmed the consistency of the association across clinically relevant strata. Calibration and decision-curve analyses further suggested acceptable risk estimation and potential clinical utility of HRR for 3-year MACE risk stratification. In the discriminative analysis for predicting 3-year MACE, the HRR had the highest area under the curve outperforming other inflammation-based indices.

**Conclusion:** A lower HRR was independently associated with a greater 3-year MACE risk in post-PCI patients. HRR outperforms commonly used leukocyte- and platelet-derived indices, highlighting its potential utility as a simple, cost-effective prognostic marker in resource-constrained healthcare settings.

## Introduction

Cardiovascular diseases (CVD) remain the leading cause of mortality worldwide^1^. According to 2021 statistics, one in three deaths globally are attributable to cardiovascular conditions^2^. This alarming figure presents a substantial challenge for healthcare systems. To address this pressing issue, advancements in medical science have been made in the areas of the diagnosis, treatment, and prognostic evaluation of CVD, particularly coronary artery disease (CAD)^3^. Among these advancements, percutaneous coronary intervention (PCI) has become a standard therapeutic approach for managing both acute and chronic CAD. The continuous development of medical technologies, along with the growing demand for effective treatments, has led to a marked increase in the number of PCI procedures performed annually^4^. This widespread adoption has contributed to a marked reduction in mortality and a clinically meaningful improvement in survival outcomes among patients with CAD^5^.

However, the increasing number of PCI cases has increased the demand for long-term patient management and follow-up, imposing a considerable burden on primary healthcare systems, particularly in the resource-limited settings of developing countries. Even in developed nations, early readmission rates among post-PCI patients remain a concern. In the United States, nearly 25% of patients who undergo PCI experience early readmission within six months of the procedure^6^. In developing countries, where primary healthcare facilities often lack diagnostic tools and specialized personnel, managing post-PCI patients becomes especially challenging. Therefore, there is an urgent need to identify simple, accessible, and cost-effective tools for monitoring and predicting outcomes in post-PCI patients.

Recent studies have highlighted the role of hematological parameters, such as leukocyte, platelet, and red blood cell indices, in the management and prognostication of CVD^7–9^. However, the use of isolated blood indices for prognostic purposes has certain limitations, particularly due to the lack of comprehensive evaluation when single indices are considered independently. To address this, composite ratios such as the neutrophil-to-lymphocyte ratio (NLR), monocyte-to-lymphocyte ratio (MLR), and systemic immune-inflammation index (SII) have gained increasing attention and have demonstrated superior prognostic value for cardiovascular outcomes^10–12^. These findings suggest that integrating multiple hematological parameters could provide more robust prognostic insights. Moreover, hematological tests are simple, low-cost, and easily accessible, making them well suited for application in primary healthcare settings with limited resources.

Among these indices, the hemoglobin-to-red cell distribution width ratio (HRR) has recently emerged as a potential prognostic tool for CVD. A retrospective study demonstrated that HRR is associated with mortality and major adverse cardiovascular events (MACE)^13^. Given its simplicity, availability, and ease of implementation, HRR could play a clinically relevant role in predicting outcomes for post-PCI patients in developing countries, providing an accessible means for follow-up in resource-limited settings. However, the HRR has not yet garnered sufficient attention in these regions, and its superiority over previously used indices has not been thoroughly evaluated.

This study aims to clarify the prognostic role of the HRR in predicting 3-year cardiovascular outcomes in post-PCI patients, particularly in the context of primary healthcare in developing countries.

## Methods

### Study design

This prospective cohort study was conducted between 01/10/2019 and 01/10/2025 at four medical centers in Vietnam: Lam Dong, Hue, Khanh Hoa, and Gia Lai. The study design incorporated elements from the Strengthening the Reporting of Observational Studies in Epidemiology (STROBE) statement to ensure the methodological rigor of the reporting. Ethical approval was obtained from the Ethics Committee of Hue University (Approval number: 918/QĐ - ĐHYH), and written informed consent was obtained from all participants. The study adhered to the principles outlined in the Declaration of Helsinki (2013 revision).

### Study population

A total of 1,019 patients who returned for their initial outpatient follow-up visit after PCI and were not readmitted during this period were initially recruited consecutively over time using a convenience sampling approach and were considered eligible for long-term follow-up. Among these patients, 703 met the inclusion criteria and were enrolled in the observational phase, while the remaining 316 met one or more exclusion criteria. Baseline data, including comorbidities, anthropometric measurements, and complete blood count parameters, were collected.

All patients received a loading dose of dual antiplatelet therapy (DAPT) in accordance with the guidelines of the European Society of Cardiology^14^. Following PCI, DAPT was continued for at least one year, after which antiplatelet monotherapy was maintained in subsequent years.

After the 3-year follow-up period, 77 patients were excluded from the final analysis due to loss to follow-up. Ultimately, 626 patients were included in the final analysis.

The exclusion criteria were as follows: contraindications to DAPT, nonadherence to prescribed antiplatelet regimens, or inability to tolerate DAPT for at least one year; concomitant serious illnesses (advanced cancer, advanced liver cirrhosis or stage IV–V chronic kidney disease); indications for cardiac surgery (severe valvular disease); life expectancy less than one year due to underlying conditions; pregnancy or intent to become pregnant; enrollment in other investigational drug or device trials not yet completed; known hematologic disorders (thalassemia, acute blood loss); and confirmed loss to follow-up (defined as failure to contact the patient or their family on at least two separate occasions, in accordance with the follow-up protocol). The detailed sampling process and recruitment sites are illustrated in **Supplementary Figure 1**.

### Clinical data collection

Baseline clinical information, including sex, age, and detailed medical history (smoking status, hypertension, diabetes mellitus, dyslipidemia, chronic kidney disease, atrial fibrillation, stroke, and heart failure), was collected at enrollment. Vital signs such as heart rate, systolic blood pressure, and body mass index (BMI) were recorded during the initial clinical examination.

For laboratory evaluations, fasting venous blood samples were obtained after a minimum of 6 hours of fasting. The complete blood count included white blood cells (WBC, G/L), neutrophils (NEU, %), lymphocytes (LYM, %), monocytes (MONO, %), eosinophils (ESO, %), red blood cells (RBC, T/L), hemoglobin (Hb, g/dL), hematocrit (HCT, %), mean corpuscular volume (MCV, fL), mean corpuscular hemoglobin (MCH, pg), mean corpuscular hemoglobin concentration (MCHC, g/dL), red cell distribution width (RDW, %), and platelet count (PLT, G/L). Additional measurements included the estimated glomerular filtration rate (eGFR) and left ventricular ejection fraction (LVEF).

The following indices were calculated on the basis of laboratory parameters: HRR = (Hb × 10) / RDW; NLR = NEU / LYM; MLR = MONO / LYM; SII = (NEU × PLT) / LYM^11,15,16^.

### Follow-up protocol and definition of study endpoints

Clinical follow-up was conducted through scheduled outpatient visits or telephone contact, including communication with family members if the patient could not be reached, in accordance with the predefined follow-up protocol. The primary endpoint was the occurrence of MACE within 3 years, defined on the basis of criteria established in previous cohort studies. Specifically, MACE in our study encompassed four components: (1) acute myocardial infarction, including new-onset myocardial infarction or coronary revascularization (either PCI or coronary artery bypass grafting); (2) hospitalization due to heart failure or deterioration in the New York Heart Association (NYHA) functional class by at least one level; (3) new-onset stroke; and (4) all-cause mortality. All MACE events were independently adjudicated by a panel of clinical experts blinded to the participants’ clinical data to ensure unbiased outcome assessment.

### Statistical analysis

All the statistical analyses were conducted via R software version 4.0.3 (R Foundation for Statistical Computing, Vienna, Austria). Missing covariates were handled using multiple imputation. Estimates were pooled using Rubin’s rules. The Kolmogorov–Smirnov test was used to assess the normality of continuous variables. Normally distributed variables are expressed as the means ± standard deviations (SDs), whereas nonnormally distributed variables are presented as medians and interquartile ranges (IQRs). Categorical variables are summarized as frequencies and percentages. As the HRR was not normally distributed, it was analyzed both as a continuous variable and as a categorical variable divided by percentile-based cutoffs. Group comparisons were performed via one-way ANOVA for normally distributed data and the Kruskal–Wallis test for nonnormally distributed data, whereas Pearson’s chi-square test was applied for categorical variables. A generalized additive model (GAM) was used to explore the potential nonlinear association between the HRR and the risk of MACE. Survival analysis was performed via Kaplan–Meier estimates, and differences in survival were assessed via the log-rank test. The associations between the HRR and MACE were examined via Cox proportional hazards regression models, with the HRR treated both as a continuous and a categorical variable. The unadjusted model included the HRR alone; Model I was adjusted for age, sex, BMI, comorbidities; and Model II, III were further adjusted for other relevant clinical and laboratory parameters. All variables included in the multivariable models were assessed for multicollinearity, with variance inflation factors below 5. Subgroup analyses were performed via stratified Cox regression, in which continuous variables were categorized on the basis of clinical thresholds or tertiles prior to interaction testing, and likelihood ratio tests were used to assess interactions. To compare the prognostic value of the HRR with that of other hematological indices, including the NLR, MLR, and SII, we constructed univariate Cox regression models for each biomarker to estimate hazard ratios (HRs), 95% confidence intervals (CIs), and p values, with model fit assessed via the Akaike information criterion (AIC). Receiver operating characteristic (ROC) curve analysis was performed to obtain the area under the curve (AUC) for each biomarker, and pairwise comparisons of AUCs were made via the DeLong test. The discriminative ability of each model was further evaluated by calculating the concordance index (C-index), and the optimal cutoff value for each biomarker was determined on the basis of the Youden index. Calibration and clinical utility were evaluated to explore the translational relevance of HRR. Calibration was assessed by comparing predicted and observed 3-year event risks derived from an HRR-based Cox model using Kaplan–Meier estimates across deciles of predicted risk. In addition, decision-curve analysis was conducted to quantify the net clinical benefit of HRR risk stratification across a range of threshold probabilities, relative to treat-all and treat-none strategies. All the statistical tests were two-tailed, with a significance level set at p < 0.05, and all the statistical reporting adhered to the Statistical Analyses and Methods in the Published Literature (SAMPL) guidelines to ensure accuracy and transparency in data presentation^17^.

## Results

### Baseline characteristics of the study population

When the anthropometric parameters and complete blood count indices of the patients across the HRR quartiles were compared, significant differences in age, sex, BMI, and hematological markers, including WBC, RBC, Hb, HCT, and RDW were detected. Additionally, heart failure with reduced ejection fraction, chronic kidney disease, and atrial fibrillation were more prevalent in the lower HRR quartiles (all p < 0.05). The detailed data are presented in **Table 1** and **Supplementary Table 1**.

**Table 1.**
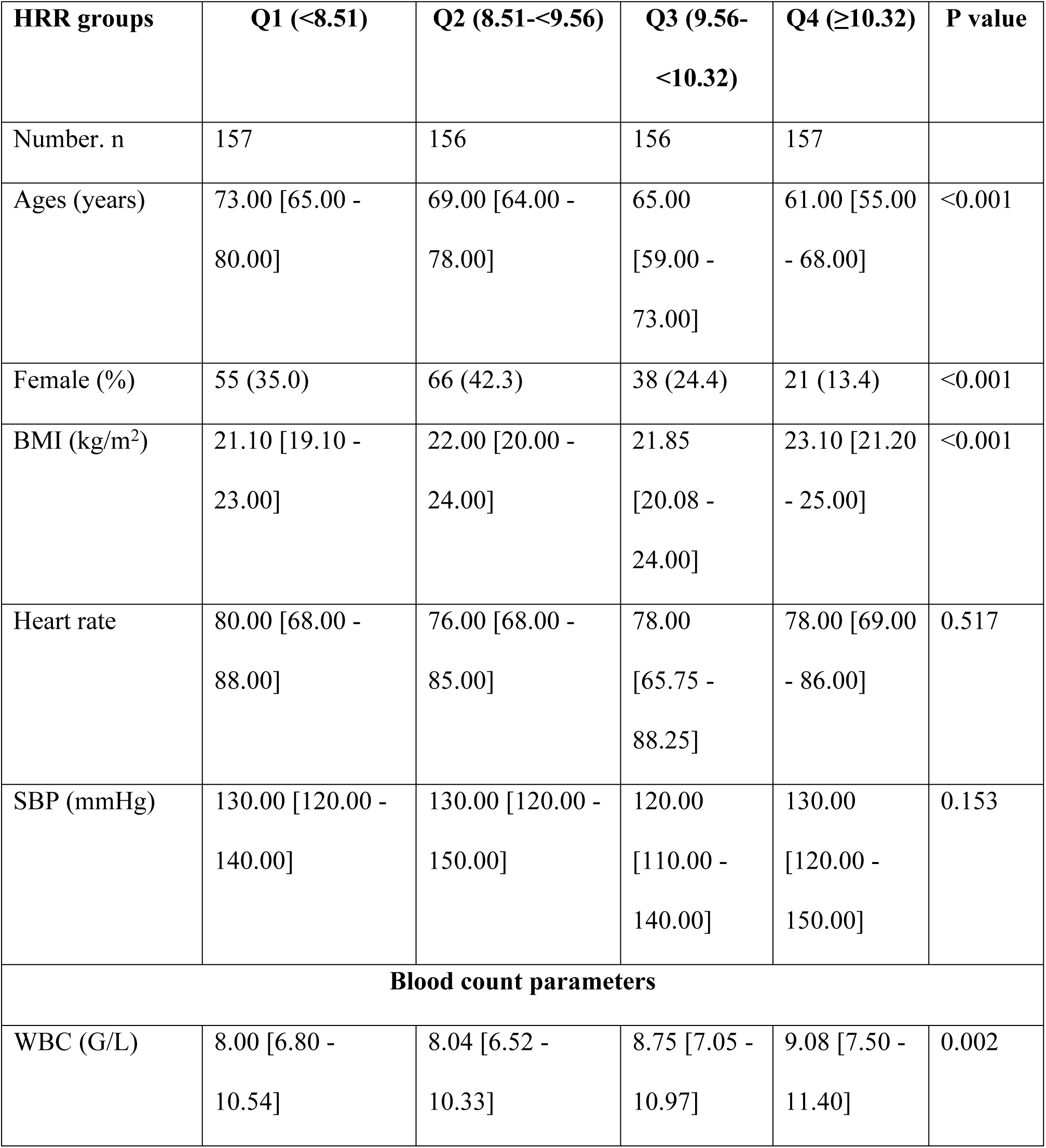
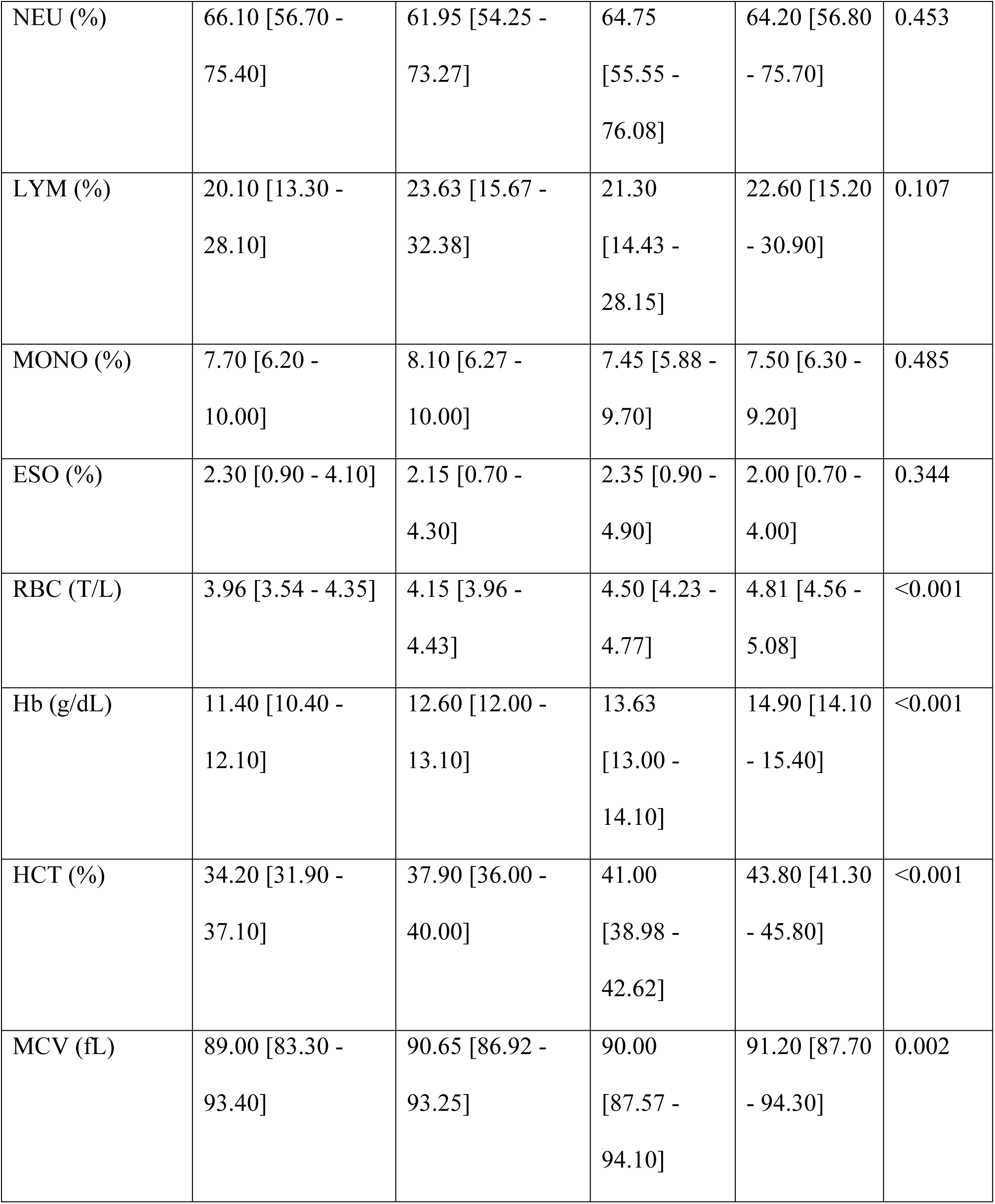
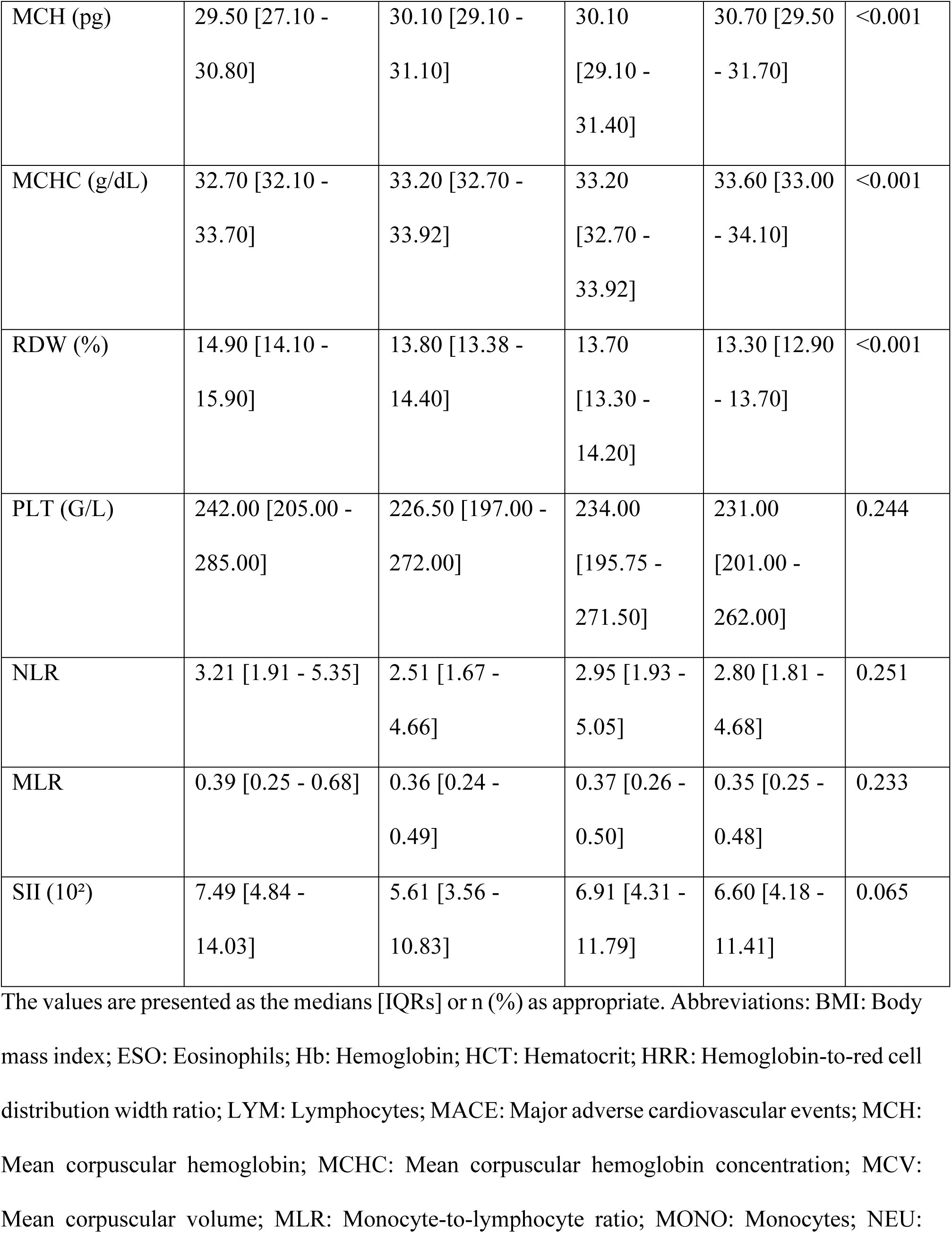
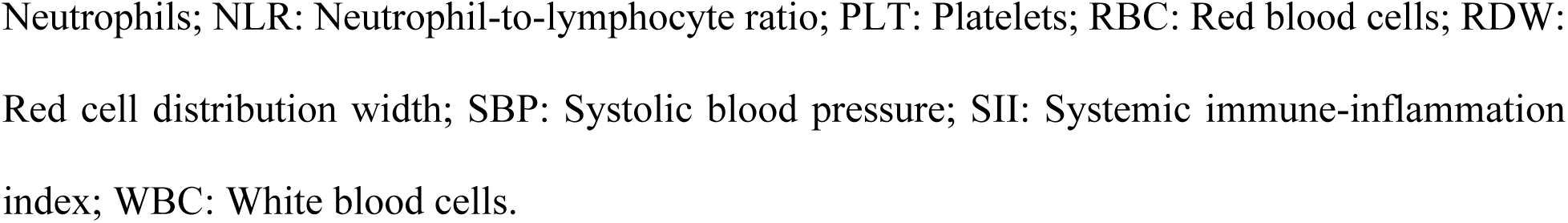
General anthropometric parameters and complete blood count indices of the patients.

### Associations between the HRR and cardiovascular events

**Figure 1** shows a progressive decline in the incidence of MACE across increasing quartiles of the HRR. Patients in the lowest HRR quartile exhibited the highest overall burden of MACE. The detailed results are presented in **Supplementary Table 2.**

**Figure 1.**
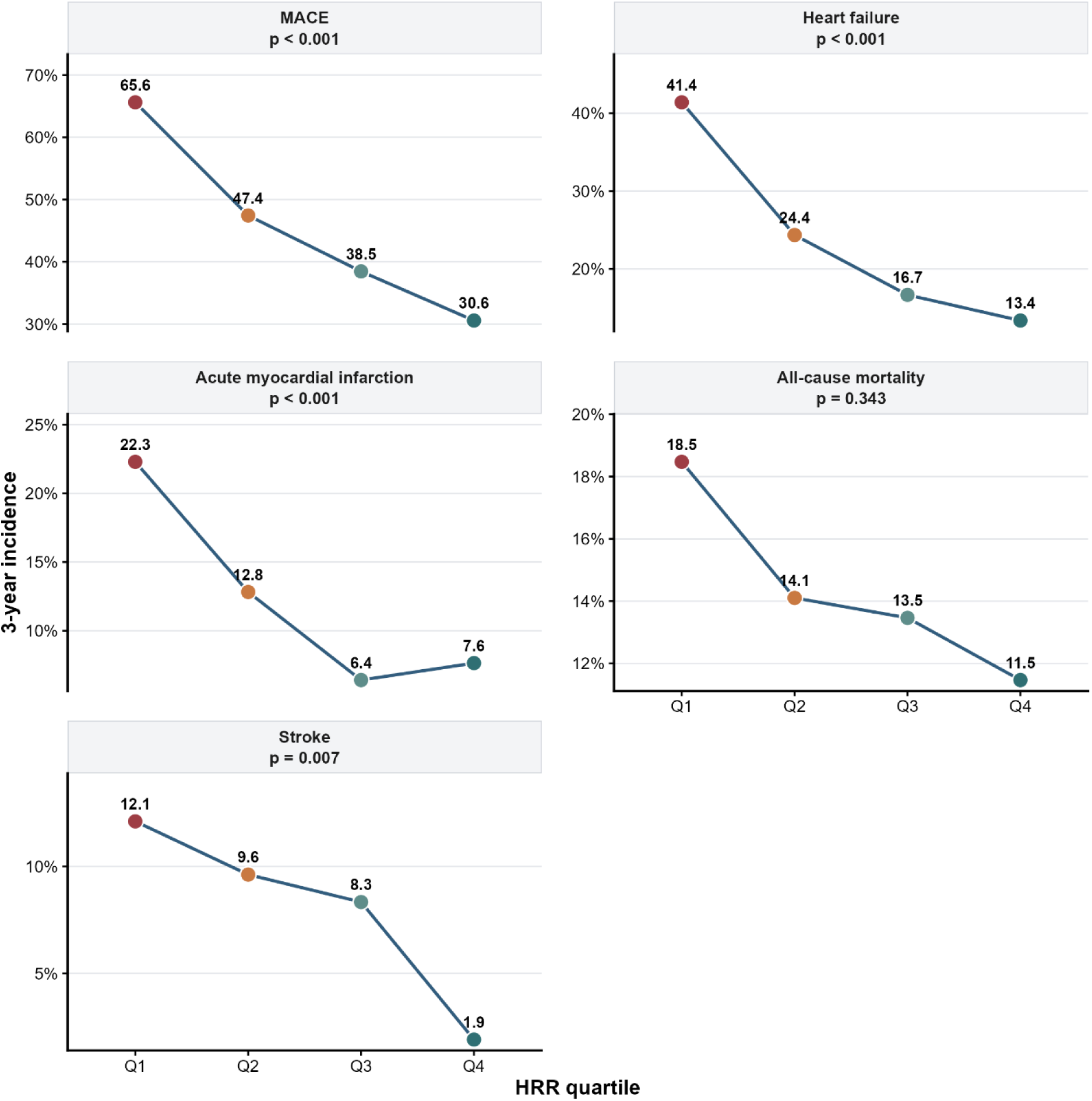
Incidence of cardiovascular events across the HRR quartiles. Abbreviations: HRR, hemoglobin-to-red cell distribution width ratio; MACE, major adverse cardiovascular events.

The nonlinear relationship between the HRR and cardiovascular outcomes demonstrated that lower HRR values were associated with a greater probability of all-cause mortality, acute myocardial infarction, heart failure, stroke, and MACE. The detailed results are illustrated in **Figure 2**.

**Figure 2.**
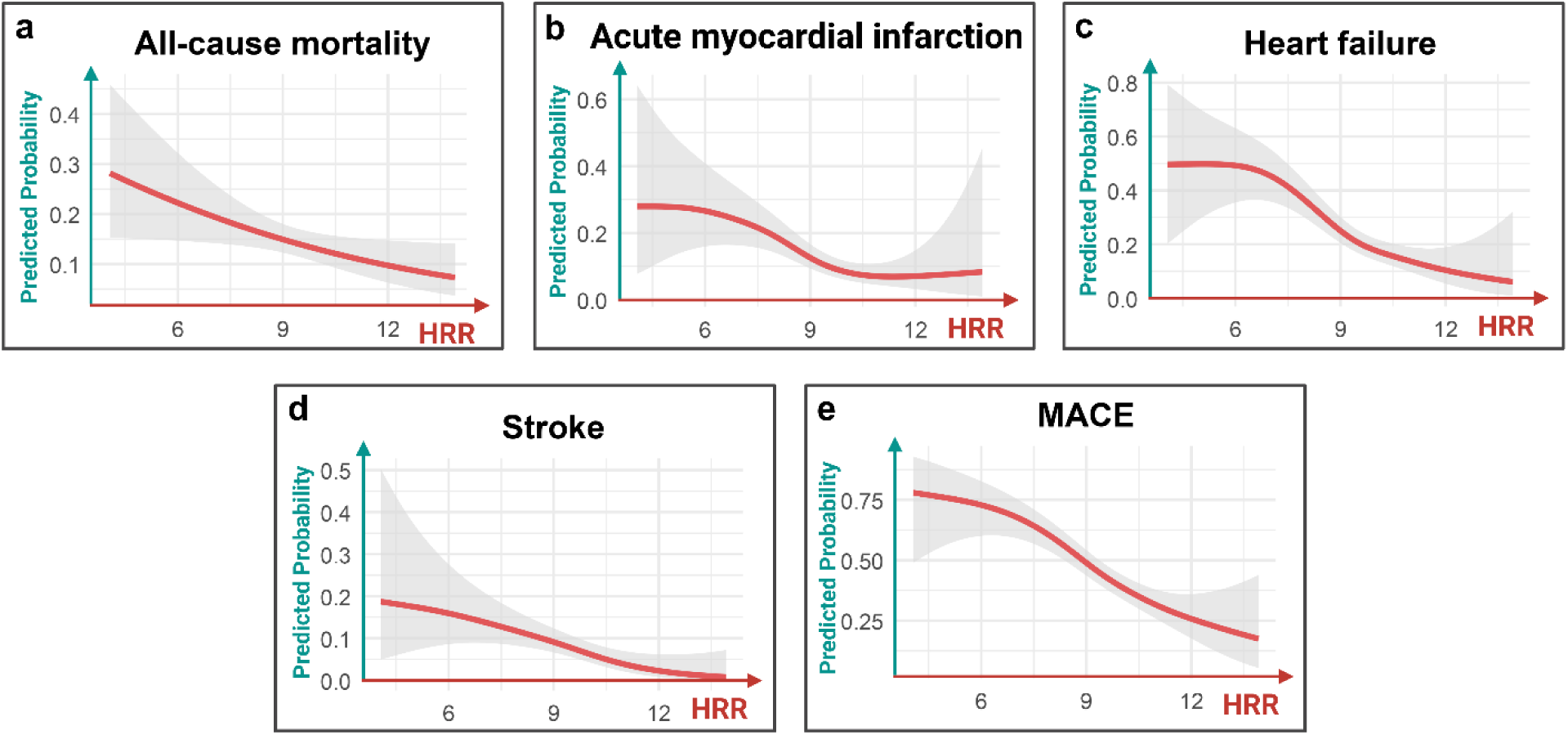
Association between the HRR and the probability of cardiovascular events based on the generalized additive model. The smooth curve fit between variables is depicted by the solid red line, whereas the 95% confidence interval is represented by dashed lines.

### Kaplan‒Meier curves of patients with different HRR values

The Kaplan–Meier curves revealed a statistically significant difference in the 3-year MACE-free rate between patients stratified by the HRR quartiles. The 3-year MACE-free rates were 34.4% for Q1, 52.6% for Q2, 61.5% for Q3, and 69.4% for Q4. A clear gradient was observed across the HRR quartiles, as illustrated in **Figure 3**.

**Figure 3.**
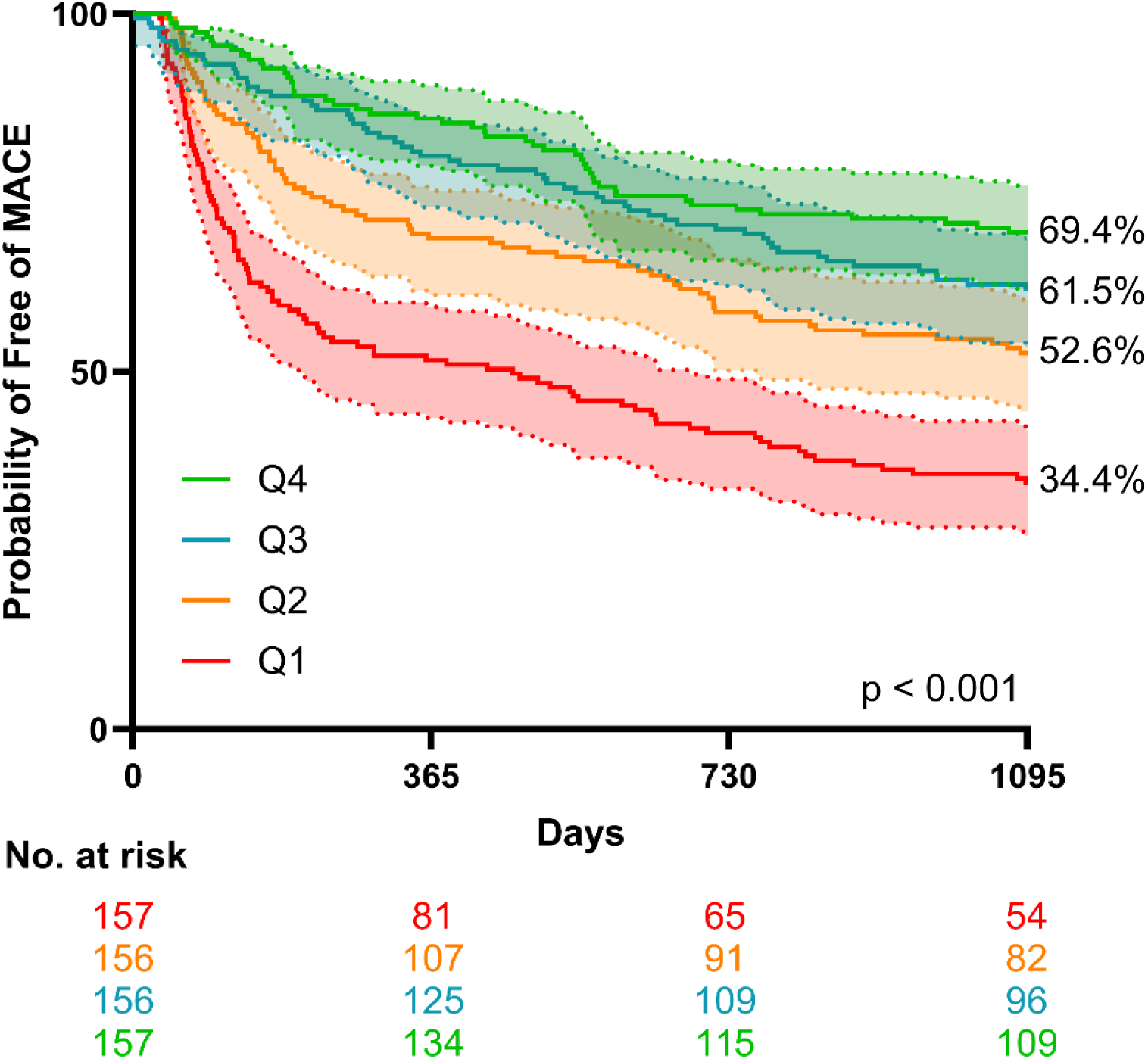
K–M curves demonstrating differences in the incidence of MACE according to the HRR group.

### Results of the adjusted and unadjusted Cox proportional hazard models

Cox proportional hazards model analysis revealed a significant inverse association between the HRR and the risk of MACE. In the unadjusted model, higher HRR was significantly associated with a lower risk of MACE (HR 0.756, 95% CI 0.703–0.814; p < 0.001). This inverse association remained significant after adjustment in Model I (HR 0.810, 95% CI 0.750–0.878; p < 0.001). Similar results were observed in the fully adjusted Model II, with HRR continuing to show a protective association with MACE (HR 0.800, 95% CI 0.738–0.868; p < 0.001). The detailed results are presented in **Table 2**.

**Table 2.**
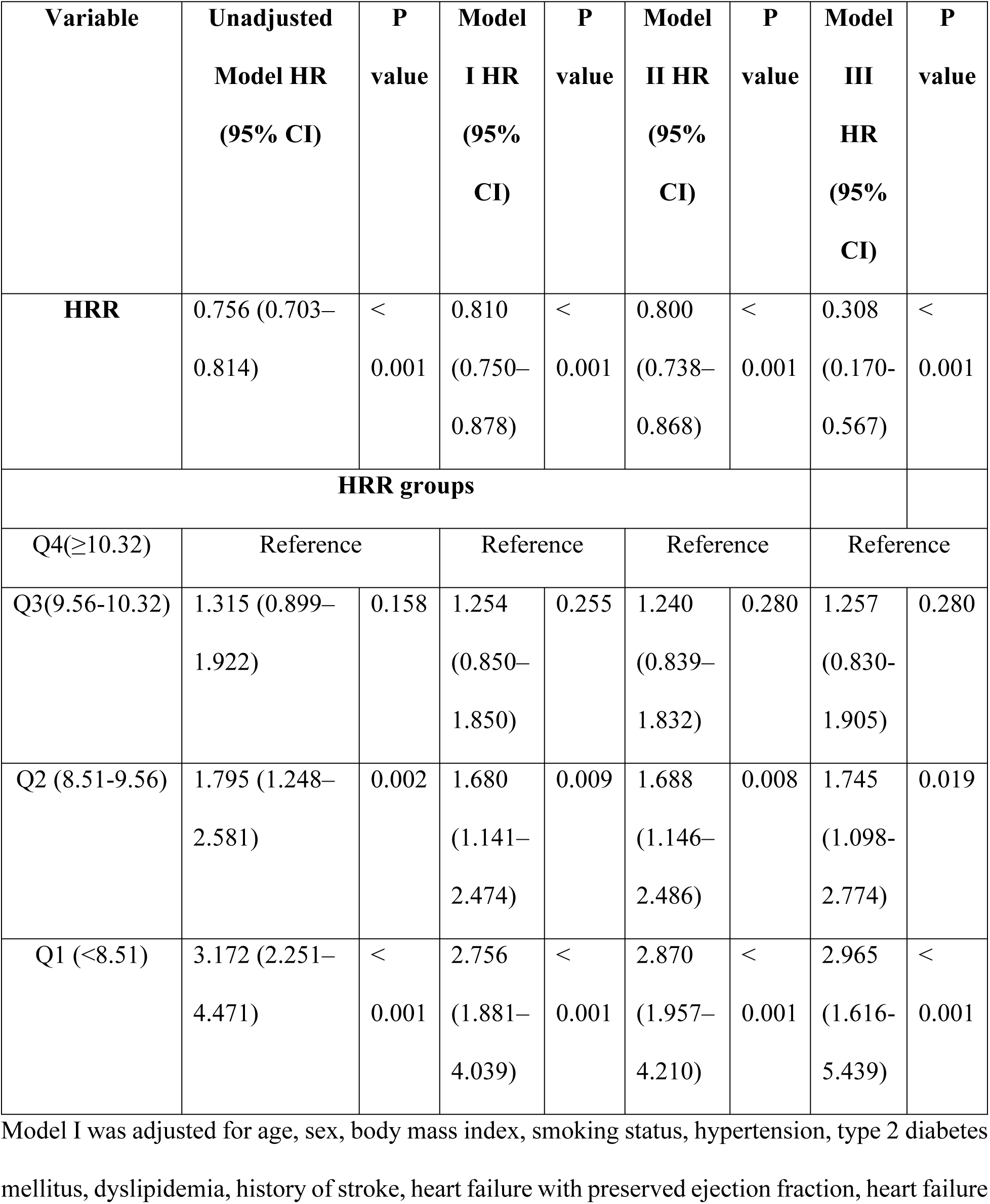
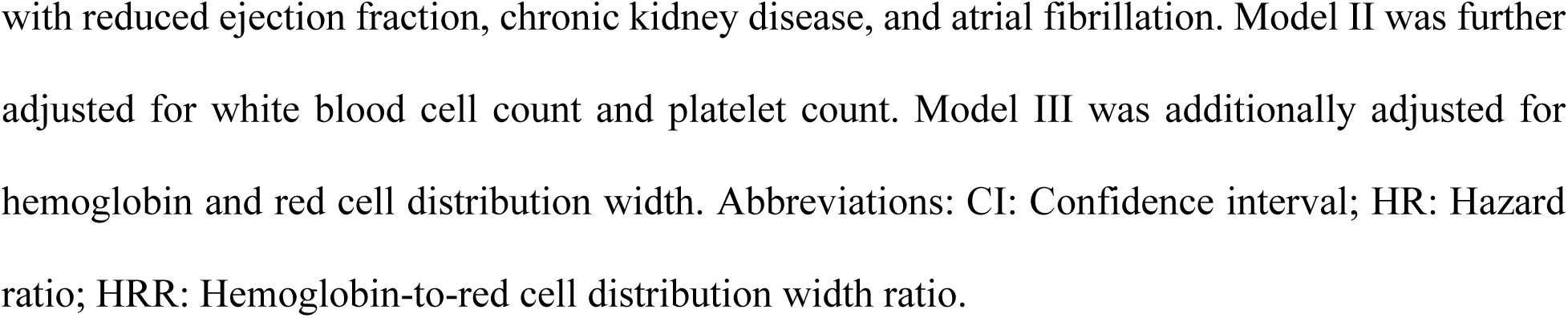
Adjusted and unadjusted Cox proportional hazards models for the associations of HRR and HRR quartiles with the risk of MACE.

| Variable | Unadjusted<br>Model HR<br>(95% CI) | P<br>value | Model<br>I HR<br>(95%<br>CI) | P<br>value | Model<br>II HR<br>(95%<br>CI) | P<br>value | Model<br>III<br>HR<br>(95%<br>CI) | P<br>value |
| --- | --- | --- | --- | --- | --- | --- | --- | --- |
| **HRR** | 0.756 (0.703–<br>0.814) | <<br>0.001 | 0.810<br>(0.750–<br>0.878) | <<br>0.001 | 0.800<br>(0.738–<br>0.868) | <<br>0.001 | 0.308<br>(0.170-0.567) | <<br>0.001 |
| **HRR groups** |  |  |  |  |  |  |  |  |
| Q4($\geq 10.32$) | Reference | | Reference | | Reference | | Reference | |
| Q3(9.56-10.32) | 1.315 (0.899–<br>1.922) | 0.158 | 1.254<br>(0.850–<br>1.850) | 0.255 | 1.240<br>(0.839–<br>1.832) | 0.280 | 1.257<br>(0.830-1.905) | 0.280 |
| Q2 (8.51-9.56) | 1.795 (1.248–<br>2.581) | 0.002 | 1.680<br>(1.141–<br>2.474) | 0.009 | 1.688<br>(1.146–<br>2.486) | 0.008 | 1.745<br>(1.098-2.774) | 0.019 |
| Q1 (<8.51) | 3.172 (2.251–<br>4.471) | <<br>0.001 | 2.756<br>(1.881–<br>4.039) | <<br>0.001 | 2.870<br>(1.957–<br>4.210) | <<br>0.001 | 2.965<br>(1.616-5.439) | <<br>0.001 |

Model I was adjusted for age, sex, body mass index, smoking status, hypertension, type 2 diabetes mellitus, dyslipidemia, history of stroke, heart failure with preserved ejection fraction, heart failure
with reduced ejection fraction, chronic kidney disease, and atrial fibrillation. Model II was further adjusted for white blood cell count and platelet count. Model III was additionally adjusted for hemoglobin and red cell distribution width. Abbreviations: CI: Confidence interval; HR: Hazard ratio; HRR: Hemoglobin-to-red cell distribution width ratio.

**Supplementary Table 3** shows that the model including HRR achieved better prognostic performance for MACE than models including Hb or RDW alone. The HRR based model demonstrated lower AIC values than the Hb- and RDW-based models, with values of 3159.6 compared with 3173.4 and 3170.5. Similarly, BIC values were lower for the HRR-based model, with values of 3207.1 compared with 3220.9 and 3217.9. In addition, the HRR-based model showed a higher C-index than models including Hb or RDW alone, with values of 0.777 compared with 0.766 and 0.775.

### Subgroup analysis

Subgroup analyses confirmed that the inverse association between HRR and 3-year MACE was consistent across all prespecified and exploratory subgroups. The findings remained robust across nearly all strata, with effect estimates that were statistically significant. These results demonstrate a high level of consistency across subpopulations. Detailed findings are presented in **Supplementary Table 4**.

### Comparison of the HRR with leukocyte- and platelet-based inflammatory indices

As shown in **Supplementary Figure 2**, the HRR demonstrated the highest discriminative ability, with an AUC of 0.649, which was significantly superior to those of the MLR (AUC = 0.571, p = 0.013), NLR (AUC = 0.557, p = 0.004), and SII (AUC = 0.559, p = 0.004). The detailed results are presented in **Supplementary Table 5**.

### Calibration and decision-curve analysis

**Supplementary Figure 3** demonstrated good agreement between predicted and observed 3-year MACE risk across low to intermediate risk ranges, indicating satisfactory calibration of the HRR-based model. At higher predicted risk levels, the model tended to slightly overestimate the observed risk, with wider confidence intervals, suggesting reduced precision in extreme risk strata. **Supplementary Figure 4** showed that the HRR-based model provided a higher net benefit than treat-all or treat-none strategies across a wide range of clinically relevant threshold probabilities (approximately 0.20–0.60), indicating meaningful clinical utility for guiding 3-year MACE risk–based decision-making.

## Discussion

To the best of our knowledge, this is the first prospective study in Vietnam to investigate the prognostic value of the HRR for long-term cardiovascular outcomes in a lower-middle-income country. In Vietnam, specialized cardiovascular services are concentrated at large tertiary hospitals, which face substantial patient overload. Strengthening primary care services, local outpatient clinics, and chronic disease management units is critical to alleviating this burden.

However, many of these peripheral healthcare settings lack access to advanced diagnostic tools, limiting their capacity to perform timely cardiovascular risk stratification and follow-up. Therefore, this study aimed to assess whether the HRR—a simple, cost-effective, and routinely available biomarker—could serve as a practical prognostic indicator in these settings.

Our findings support the clinical utility of the HRR, demonstrating that lower HRR values are significantly associated with a greater risk of 3-year MACE in patients undergoing PCI. These results are consistent with those of retrospective studies conducted in China, which revealed that a lower HRR was predictive of increased post-PCI mortality among patients with CAD. For example, a study by Zhang et al. revealed significantly greater mortality in the low HRR group than in the high HRR group^13^.

Previous evidence has revealed a strong association between low Hb levels and increased mortality in patients with CAD^18,19^. Anemia is also known to be independently associated with a greater risk of heart failure hospitalization and all-cause mortality in patients undergoing coronary angiography^20,21^. Moreover, secondary analyses of the SPRINT trial and large cohorts such as the ARIC confirmed anemia as an independent risk factor for adverse cardiovascular outcomes and death^22,23^. Anemia is an independent risk factor for CVD outcomes in the ARIC cohort, a community cohort of subjects between the ages of 45 and 64 years^23^. A meta-analysis further demonstrated that anemia significantly worsens the prognosis of acute heart failure, predicting both short- and long-term all-cause mortality and heart failure events^24^.

On the other hand, elevated RDW has also been identified as a predictor of mortality in patients undergoing PCI. A previous study demonstrated that higher RDW is associated with greater myocardial injury in patients with non-ST-elevation ACS^25^. In addition, RDW levels are more elevated in patients with heart failure—particularly those with acute heart failure—compared to those with chronic heart failure^26–28^. Post hoc analyses from trials such as ODYSSEY OUTCOMES further support that RDW is independently associated with MACE and death in stable post-ACS patients^29^.

Evidence indicating that decreased Hb and elevated RDW are associated with a higher risk of MACE provides an indirect rationale for the prognostic relevance of HRR, which is calculated as the ratio of Hb to RDW. Accordingly, patients with lower HRR values may be at greater risk of experiencing adverse cardiovascular events. However, it remains unclear whether HRR provides prognostic information beyond its individual components, Hb and RDW. In a sensitivity analysis, the association between HRR and 3-year MACE remained statistically significant after additional adjustment for Hb and RDW.

In addition to the HRR, inflammatory indices derived from complete blood counts—such as the NLR and SII—also demonstrated prognostic value in our study. These findings align with prior research: the PARIS study reported that an elevated WBC count was an independent predictor of MACE after PCI^30^. Similarly, higher pretreatment NLR values have been associated with increased risks of MACE and mortality in patients with acute coronary syndrome^15^. SII has also been linked to adverse cardiovascular outcomes in individuals undergoing PCI^16^. However, when directly compared, HRR demonstrated the highest AUC among these indices for predicting 3-year MACE.

The HRR showed good agreement between predicted and observed 3-year MACE risk across low-to intermediate-risk ranges, indicating acceptable calibration. In addition, decision-curve analysis demonstrated a positive net benefit across clinically relevant threshold probabilities, suggesting that HRR may be useful for risk stratification rather than direct therapeutic decision-making.

Importantly, the ability to calculate the HRR and other inflammatory markers from routine complete blood counts has substantial clinical value, especially in resource-limited settings. These biomarkers are simple, inexpensive, and feasible for widespread use at the primary care level. HRR, in particular, should be considered a supportive tool alongside traditional clinical assessments, laboratory evaluations, and established risk scores to enable more accurate and comprehensive risk stratification, thereby improving patient care and outcomes.

## Limitations

Despite the strengths of our prospective, multicenter study design, several limitations should be acknowledged. First, the study population was limited to patients in Vietnam, which may affect the generalizability of the findings to other geographic or ethnic populations. External validation in diverse settings is warranted. Second, although all-cause mortality was chosen as a robust and unbiased endpoint, the study did not differentiate between cardiovascular and noncardiovascular causes of death. This approach was necessitated by the challenges in accurately determining causes of death in Vietnam, where autopsy and systematic cause-of-death classification are not routinely performed owing to cultural, religious, and logistical constraints. Third, excluding patients readmitted before first outpatient follow-up and those unable to maintain DAPT for at least one year likely removes higher-risk individuals, inflating apparent prognostic performance and limiting generalizability. Fourth, blood testing procedures were not fully standardized across different participating centers, which may have introduced inter-laboratory variability. Fifth, inflammatory and nutritional markers, such as CRP, ferritin, vitamin B12, folate, and IL-6, were not routinely measured because these tests are not consistently available in resource-limited primary healthcare settings. Therefore, their potential influence on HRR and any effect modification of the association between HRR and clinical outcomes could not be assessed.

## Conclusions

A lower HRR was significantly associated with an increased risk of 3-year MACE in patients undergoing PCI. HRR was an independent predictor of adverse cardiovascular outcomes, outperforming several commonly used inflammation-based indices.

## DATA AVAILABILITY STATEMENT

Research data supporting this publication will be available through direct request to the first author Hai Nguyen Ngoc Dang

## AUTHOR CONTRIBUTIONS

Hai Nguyen Ngoc Dang: Conceptualization, methodology, investigation, data curation, writing – original draft, writing – review & editing. Thang Viet Luong: Investigation, Data curation, Writing – original draft, Writing – review & editing. Binh Anh Ho, Tien Anh Hoang, Minh Van Huynh: supervison. Thanh Thien Tran, Thanh Van Ho, Mai Thi Thu Cao, Tan Ngoc Nguyen, Binh Anh Ho, Minh Van Huynh, Khiem Dong Thien, Cuong Hai Nguyen, Hung Nguyen Minh: Writing – original draft, Writing – review & editing. All the authors have read and agreed to the published version of the manuscript.

## ETHICS APPROVAL STATEMENT

The research was conducted following the guidelines stipulated in the Helsinki Declaration. Our research was approved by the Institutional Ethics Committee of Hue University (No: 918/QĐ - ĐHYH).

